# Relation of spice consumption with COVID-19 first wave statistics (infection, recovery and mortality) across India

**DOI:** 10.1101/2022.06.08.22275684

**Authors:** Vedvati Bhapkar, Supriya Bhalerao

## Abstract

**Background & Objectives:** The recovery and mortality statistics for COVID-19 first wave considerably differed in different states & Union territories (UT) of India. Though dependent on several factors, relation of diet and immunity is well-established. Spices are an essential part of Indian cuisine. Apart from adding flavors and colors to the food, their importance has been traditionally known in disease prevention and cure. Thus, present study was carried out to assess relation of spice consumption with COVID-19 first wave statistics in India.

**Methods:** The spice consumption data were retrieved from ‘Household Consumption of Various Goods and Services in India’ from 68^th^ round (2011-12) of survey conducted by National Sample Survey Organization (NSSO). Spices for which, consumption data was available, viz., ginger (*Zingiber officinale*), garlic (*Allium sativum*), cumin (*Cuminum cyminum*), coriander (*Coriandrum sativum*), turmeric (*Curcuma longa*), black pepper (*Piper nigrum*), chili (*Capsicum annuuam*), tamarind (*Tamarandus indica*) and ‘other spices’ were selected for analysis. The COVID-19 first wave data for individual states and UTs were retrieved as total number of cases, number of cured/discharged/migrated cases and total number of deaths due to COVID-19, in a cumulative form. It was normalized ‘per million’ population of respective states and UT. The correlation of individual spice consumption and COVID-19 statistics was analyzed.

**Results and Conclusions:** Spices were consumed across all India with a varied range. The highest consumed spice was ginger. Its highest consumption was in Mizoram (185 gm/30 days) and least in Jammu & Kashmir (23gm/30 days). The highest consumption of ‘Other spices’ were observed in Lakshadweep (149 gm/30 days), which incidentally reported zero COVID-19 cases. Tamarind consumption showed positive correlation (r = 0.4724) with total number of cases per million population, recovered/migrated/cured cases (r = 0.4948). The consumption of cumin exhibited a weak positive correlation (r = 0.5011) with total deaths per million population. However, most of these correlations were statistically insignificant. The findings from this study provide a basic framework and understanding for future studies. These findings can help to predict preventive/ mitigating or curative usage of these spices. Should similar scenario occur in future, these findings can provide some vital base to act as adjuvant management. As the unspecified and under-explored ‘Other spices’ category showed promising correlation, more attention needs to be given to them too, along with mostly studied spices like ginger and turmeric.

## Introduction

The COVID-19 first wave in India started around January 2020 and continued for almost a year.[1] During this wave, there was a great display of variation in its incidence in different parts of the world. India, the second largest populated nation, was second in COVID-19 case tally for most of the time.[2] However, the incidence as well as mortality due to COVID-19 in India per one million population was considerably lower than the USA and Brazil, which were at first and third position then.[3] Like the global scenario, it was observed that not all regions of India were equally affected. The recovery and mortality statistics considerably differed in different states & Union territories (UT) of India.[4] It has been proposed that the immune system plays a major role in etiopathogenesis of COVID-19.[5] Though dependent on several factors, relation of diet and immunity is well-established.[6] Spices are a distinctive feature of Indian diet. They are cultivated and consumed all over India as per climatic variations. Spices basically add flavor and colors to the food. Also, their importance has been traditionally known in prevention and cure of different ailments.[7] A few in silico studies have reported that some chemical constituents from Indian spices are significantly active against COVID-19. Phytochemicals from black pepper, ginger, and garlic are successfully studied for their COVID-19 management potential. [8] ‘Curcumin’, an alkaloid obtained from turmeric holds similar promising results. [9] Also, certain chemical compounds isolated from Indian spices were recommended as potential drug candidates against COVID-19. [10] ‘Spicerx’, an extensive evidence-based database regarding spices, their constituents and health benefits has also been recently compiled.[11] Elsayed & Khan have analyzed COVID-19 records for first wave from 163 countries and revealed a clear correlation of its incidence, recovery rate and mortality rate with their spice consumption.[11] They have reported that total number of cases and mortality were lower in countries with high spice consumption. The recovery rate was also higher in these countries. As Indian cuisine is famous for its spiciness, we decided to carry out the relation of spice consumption with COVID-19 statistics, during first wave in India. To the best of our knowledge, no such study has been carried out in India till date. Simple dietary items are mostly used as household remedies here. It was interesting to explore if, spices, which are an integral yet modest part of Indian cuisine can also help in managing deadly diseases like COVID-19. Further, it was thought interesting in view of huge diversity in epidemiology of various diseases [12] as well as variations in meal pattern [13] amongst the different states of India.

## Methodology

The spice consumption data were retrieved from ‘Household Consumption of Various Goods and Services in India’ from 68th round (2011-12) of consumption expenditure survey conducted by National Sample Survey Organization (NSSO).[14] Spices for which, consumption data was available, viz., ginger (*Zingiber officinale*), garlic (*Allium sativum*), cumin (*Cuminum cyminum*), coriander (*Coriandrum sativum*), turmeric (*Curcuma longa*), black pepper (*Piper nigrum*), chili (*Capsicum annuuam*) and tamarind (*Tamarandus indica*) were selected for further analysis. The category mentioned as ‘other spices’ was also selected as it may incorporate spices specifically used in that particular state or UT. There was no mention in the survey document regarding which spices were included in this ‘other spices’ category. In the survey report, rural and urban consumption of spices was presented separately. An average of these two was taken to get an overall consumption record of each spice category. The consumption of each of these nine categories, consumed in gram, over a period of 30 days was tabulated.

The COVID-19 data for individual states and UT of India were retrieved from the website of Ministry of Health & Family Welfare, Govt. of India, on 25^th^ September 2020. The data were obtained as total number of cases, number of cured/discharged/migrated cases and total number of deaths due to COVID-19, in a cumulative form. It was normalized ‘per million’ population of respective states and UT, on the basis of their populations obtained from official census data for further analysis.[15] The fractions were converted to absolute numbers.

The Telangana state was formed by separating from Andhra Pradesh in 2014. The Union territory of Ladakh was formed in 2019 from Jammu & Kashmir state. Both these were not separately existent in 2012 when this NSSO survey was carried out. Hence, their COVID-19 statistics were merged with their respective parent states. The COVID-19 statistics for UTs of Dadara & Nagar Haveli and Daman & Diu were available collectively. Hence, their spice consumption data were averaged out.

Scatter charts were plotted in MS-Excel to observe correlation between individual spice consumption and total cases of COVID-19, recovered cases and mortality per million populations. Spearman’s rank test was applied to analyze this correlation using GraphPad InStat software version 3.5. The significance of correlation was fixed at p<0.05.

## Results

The state wise data of first wave COVID-19 statistics and average spice consumption is mentioned in Table 1. It was seen that ginger, garlic, cumin, coriander, turmeric, black pepper, chili and tamarind are consumed across all India but in varying amounts. The range of average consumption largely varied for all these spices. The ranges of consumption in gm/30 days for individual spices were ginger (23-185), garlic (50-168), cumin (1-77), coriander (2-99), turmeric (10-91), black pepper (0-27), chili (7-161) and tamarind (0-225). Other spices were consumed in the range of 18-149 gm per 30 days. The highest ginger consumption/30 days was observed in Mizoram (185 gm). Pondicherry showed highest consumption of garlic (168 gm). The consumption of cumin was highest in Chandigarh (77 gm) and lowest in Mizoram & Nagaland (each 1 gm). Coriander was most consumed in Kerala (99 gm) and least in Mizoram (2 gm). Consumption of turmeric was highest in Jammu & Kashmir (including Ladakh) as 91 gm and lowest in Nagaland with only 10 gm. Black pepper was consumed in very less quantities amongst all these spices. Tamil Nadu (27 gm) showed highest consumption of black pepper and it was zero for Mizoram, Nagaland and Sikkim. Chilies were most consumed in Kerala (161 gm), closely followed by Lakshadweep (153 gm). Least consumption of chilies was seen in Meghalaya (7 gm). Tamarind was mostly consumed in large quantity in southern states and UTs. Largest tamarind consumption was seen in Pondicherry (225 gm). Many states such as Himachal Pradesh, Meghalaya, Nagaland, Sikkim, Tripura, Uttar Pradesh and Uttarakhand even reported zero tamarind consumption. The highest consumption of ‘other spices’ were observed in Lakshadweep (149 gm) followed by Andaman & Nicobar Islands (72 gm). Looking at the COVID-19 first wave scenario, highest number of cases per million population were found in Goa (19713), followed by Pondicherry (18583). However, the number of recovered cases was also substantial for them. Lakshadweep displayed zero incidences. Mizoram considerably had zero mortality and only 1542 cases per million population. Madhya Pradesh had 1489 cases per million population and mortality of only 28 per million population. The combined mortality of UTs Dadara & Nagar Haveli and Daman & Diu was as low as three cases per million population. The scatter graphs for correlation of state-wise first wave COVID-19 statistics with spice consumption is presented in Figure 1. When correlation between spice consumption and total cases of COVID-19, recovered cases and mortality per million population was analyzed, we observed a negative correlation between consumption of ginger with total number of cases as well as number of deaths per million population. Turmeric also showed negative correlation with number of deaths per million population. However, both these correlations were statistically insignificant. The number of recovered cases had a positive correlation with consumption of all spices except ginger and turmeric. However, it was significant in case of tamarind and other spice consumption only.

**Table 1:** State wise data of first wave COVID-19 statistics and average spice consumption.

| State/UT | Average Spice consumption (gm/ 30 days) |  |  |  |  |  |  |  |  | COVID-19 data (per million population) |  |  |
| --- | --- | --- | --- | --- | --- | --- | --- | --- | --- | --- | --- | --- |
|  | Ginger | Garlic | Cumin | Coriander | Turmeric | Black Pepper | Chili | Tamarind | Other spices | Total cases | Recovered or migrated cases | Deaths |
| Andaman & Nicobar | 80 | 92 | 32 | 45 | 42 | 9 | 68 | 90 | 72 | 9651 | 9115 | 137 |
| Andhra Pradesh | 83 | 87 | 34 | 62 | 44 | 3 | 124 | 173 | 38 | 17736 | 15247 | 139 |
| Arunachal Pradesh | 120 | 76 | 19 | 27 | 32 | 4 | 30 | 3 | 47 | 5489 | 4078 | 9 |
| Assam | 85 | 77 | 26 | 16 | 57 | 3 | 21 | 1 | 22 | 5105 | 4138 | 19 |
| Bihar | 28 | 65 | 25 | 58 | 60 | 15 | 45 | 1 | 26 | 1630 | 1501 | 8 |
| Chandigarh | 78 | 52 | 77 | 61 | 87 | 7 | 76 | 2 | 40 | 9757 | 7022 | 119 |
| Chhattisgarh | 62 | 69 | 18 | 60 | 54 | 2 | 47 | 8 | 32 | 3452 | 1940 | 27 |
| Dadara& Nagar Haveli, Daman & Diu | 96 | 79 | 25 | 42 | 40 | 5 | 72 | 15 | 46 | 5007 | 4628 | 3 |
| Delhi | 88 | 53 | 39 | 45 | 44 | 12 | 45 | 1 | 39 | 14848 | 12706 | 299 |
| Goa | 63 | 73 | 25 | 47 | 36 | 5 | 107 | 34 | 62 | 19713 | 15581 | 247 |
| Gujrat | 73 | 85 | 36 | 54 | 46 | 9 | 93 | 10 | 43 | 2062 | 1737 | 55 |
| Haryana | 66 | 64 | 46 | 45 | 66 | 3 | 61 | 1 | 33 | 4460 | 3585 | 46 |
| Himachal Pradesh | 67 | 72 | 39 | 48 | 65 | 7 | 21 | 0 | 36 | 1812 | 1144 | 19 |
| Jammu & Kashmir | 23 | 72 | 12 | 8 | 91 | 2 | 94 | 8 | 45 | 19286 | 13415 | 266 |
| Jharkhand | 53 | 74 | 28 | 32 | 51 | 16 | 24 | 2 | 18 | 2203 | 1775 | 19 |
| Karnataka | 53 | 94 | 33 | 67 | 33 | 16 | 119 | 66 | 22 | 8624 | 6930 | 133 |
| Kerala | 72 | 65 | 21 | 99 | 35 | 18 | 161 | 98 | 64 | 4150 | 2955 | 17 |
| Lakshadweep | 89 | 67 | 24 | 88 | 34 | 19 | 153 | 47 | 149 | 0 | 0 | 0 |
| Madhya Pradesh | 65 | 68 | 33 | 90 | 43 | 3 | 95 | 1 | 29 | 1489 | 1151 | 28 |
| Maharashtra | 69 | 109 | 35 | 36 | 42 | 6 | 97 | 4 | 47 | 10896 | 8154 | 294 |
| Manipur | 121 | 73 | 8 | 17 | 40 | 1 | 76 | 1 | 39 | 3505 | 2660 | 23 |
| Meghalaya | 112 | 74 | 5 | 15 | 33 | 4 | 7 | 0 | 19 | 1595 | 852 | 12 |
| Mizoram | 185 | 69 | 1 | 2 | 52 | 0 | 18 | 1 | 29 | 1542 | 922 | 0 |
| Nagaland | 144 | 92 | 1 | 5 | 10 | 0 | 75 | 0 | 28 | 2802 | 2263 | 8 |
| Odisha | 65 | 68 | 28 | 11 | 39 | 1 | 32 | 18 | 27 | 4387 | 3559 | 17 |
| Pondicherry | 64 | 168 | 30 | 98 | 29 | 26 | 113 | 225 | 49 | 18583 | 14476 | 374 |
| Punjab | 89 | 61 | 26 | 27 | 76 | 6 | 56 | 3 | 56 | 3602 | 2718 | 103 |
| Rajasthan | 52 | 96 | 35 | 82 | 48 | 4 | 137 | 1 | 21 | 1705 | 1419 | 20 |
| Sikkim | 142 | 85 | 12 | 13 | 32 | 0 | 21 | 0 | 19 | 4140 | 3253 | 47 |
| Tamil Nadu | 54 | 95 | 33 | 90 | 30 | 27 | 99 | 138 | 50 | 7586 | 6819 | 123 |
| Tripura | 57 | 50 | 33 | 20 | 51 | 1 | 32 | 0 | 34 | 6215 | 4323 | 68 |
| Uttar Pradesh | 57 | 66 | 25 | 63 | 45 | 5 | 38 | 0 | 40 | 1796 | 1449 | 26 |
| Uttarakhand | 66 | 70 | 32 | 90 | 44 | 2 | 44 | 0 | 22 | 4142 | 2895 | 50 |
| West Bengal | 99 | 53 | 38 | 15 | 58 | 3 | 27 | 3 | 45 | 2501 | 2180 | 48 |

**Figure 1:**
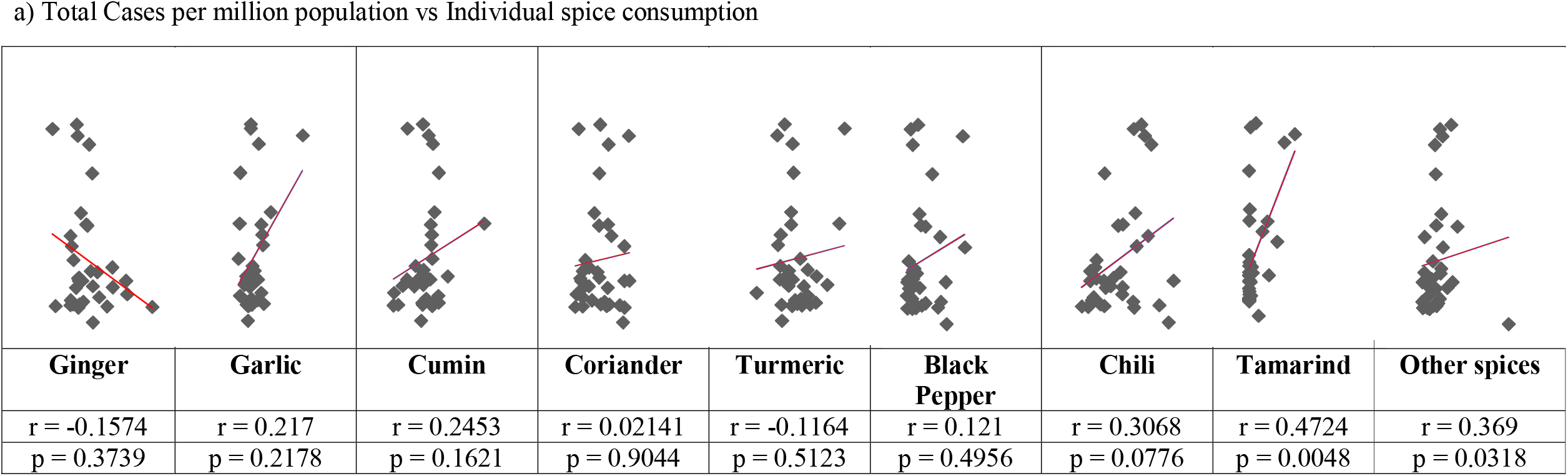
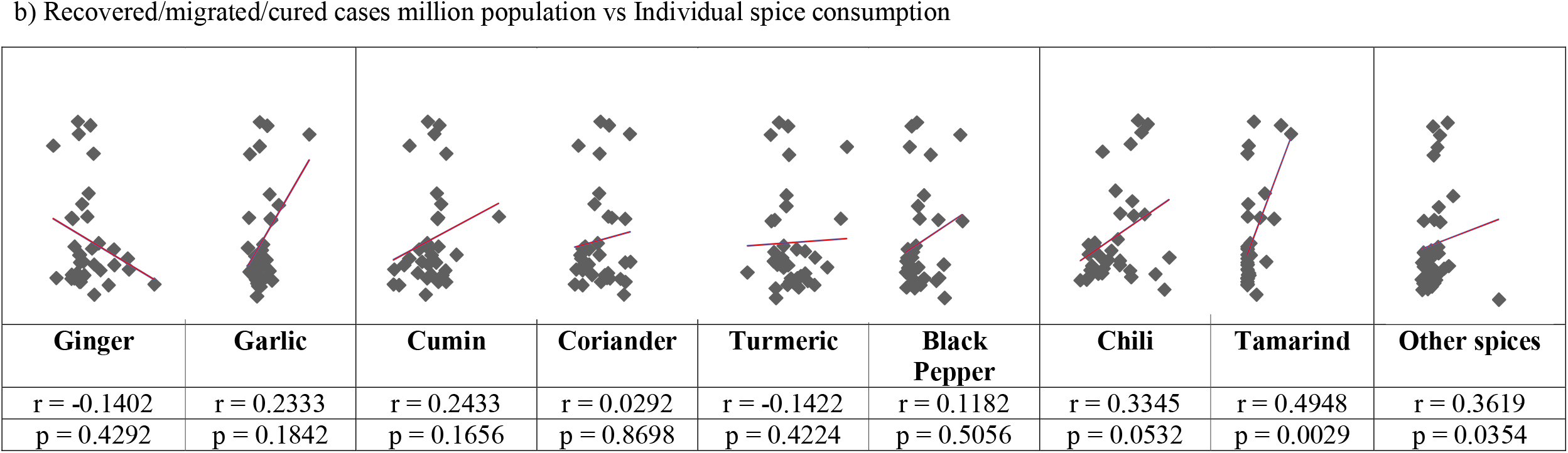
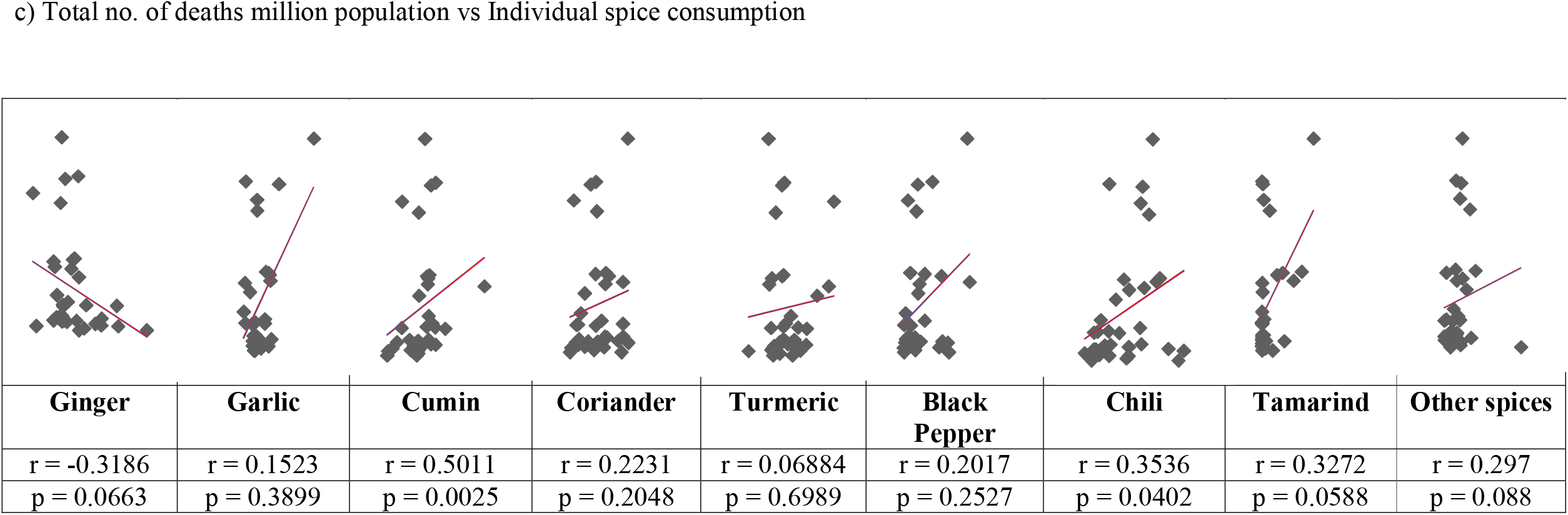
Correlation of state-wise first wave COVID-19 statistics with spice consumption (X-axis-Consumption in gm/30 days; Y-axis-no. of cases/ million population)

## Discussion

The present study was carried out to assess relation between spice consumption and COVID-19 first wave statistics. It revealed that the consumption of chili, tamarind and spices listed under the category ‘other spices’ had significant positive correlation with the number of recovered cases during first wave. Many studies are carried out for majorly consumed spices such as ginger, turmeric, garlic etc. The overlooked category of ‘Other spices’ showed promising correlation and thus needs further focus too.

Spices are food adjuncts that have taste-enhancing as well as medicinal effects. Along with traditional knowledge, various scientific studies support medicinal potential of spices. In current situation, the spice herbs are being looked up to for their potential action against COVID-19. Host immune response and host inflammatory response are vital in prevention and recovery from COVID-19.[16] The spices included in the present study have shown a promise in this regard. Ginger has shown anti-inflammatory [17] as well as immunomodulator [18] properties in experimental studies. The action of garlic on cytokine secretion and thus immunomodulatory and anti-inflammatory activities have been established.[19] Cumin oil has shown anti-inflammatory action through inhibiting NF-_κ_B and mitogen activated protein kinases as well as immunomodulatory action.[20] Various studies have suggested that coriander and its bioactive constituents possess anti-inflammatory properties.[21] Curcumin, derived from turmeric has been extensively proven to demonstrate anti-inflammatory and immunomodulatory activities.[22] The essential oils from black pepper have shown anti-inflammatory properties.[23] Its potential immunomodulatory activity has also been documented.[24] The capsaicinoids in chili have also shown anti-inflammatory properties.[25] The fruit pulp of tamarind has shown immunomodulatory activities.[26] As mentioned earlier, some spices and their actives have also been shown to be effective against COVID-19 infection.

Elsayed and Khan had shown a correlation between high consumption of spices with low infection and mortality due to COVID-19 at global level. Their study also suggested a probable role of spices against COVID-19. However, their data did not clearly indicate individual spices incorporated in study. Our study shows relationship between consumption of specific spices with COVID-19 incidence, mortality and recovery across states and UTs of India, during the first wave.

As per The Spices Board of India, 52 spices are grown and consumed in India.[27] The spices considered for present study, viz. ginger, garlic, cumin, coriander, turmeric, black pepper, chili and tamarind are most commonly used in culinary practices across India. Apart from them, certain spices such as cardamom, cinnamon, asafetida, nutmeg etc. are used but in lesser quantities and as per local cuisine preference. Thus, the category of ‘Other spices’ was also taken into consideration to get an inclusive consumption data.

We observed that the range of spice consumption varied greatly throughout the country. Maximum range was observed for tamarind. Its consumption was almost limited to southern part of India (225 gm/30 days), while in the North East India, its consumption is almost nil. The UT of Lakshadweep showed highest consumption of ‘other spices.’ It is important to explore this category further as Lakshadweep had zero incidence of COVID-19. It was observed that spices such as curry leaves, cardamom, cloves etc. were used more in Lakshadweep cuisine. [28] These can be explored further for their anti-COVID-19 potential.

Interestingly, ginger consumption showed a negative correlation with incidence, mortality as well as recovery from COVID-19. On the contrary, garlic showed a positive correlation with them. Thus, ginger may have a role in prevention of COVID-19 and garlic in its recovery. Also, turmeric had a negative correlation with incidence of COVID-19. Thus, ginger and turmeric may have immune potentiating property. Also, all other spices that exhibited positive correlation with recovery from COVID-19 may possess anti-viral properties. It should be noted here that spices are consumed in very less quantities unlike other dietary components such as cereals, legumes, vegetables and meat. Although, their stand-alone consumption analysis may imply non-significant correlations, spices may be vital in a bigger picture collectively with other food components.

The spice palate of India is shared by its neighboring countries, viz. Pakistan, Bangladesh, Nepal and Sri Lanka. As their diet consists of more animal sourced components such as meats, fishes etc.; it can be understood that these spices get consumed in larger quantities over there. Remarkably, the incidence as well as mortality due to COVID-19 was very low in these countries compared to India, during first wave.

A major limitation of our study is unavailability of recent official data for consumption of the spices. Though a nation-wide survey was carried out in 2017-18; its results were not published on the NSSO website.[29] It is possible that consumption patterns may have changed owing to a time lapse of almost ten years. The exact components of ‘Other spices’ category couldn’t be retrieved from NSSO website. It would be a good idea to collect the data about consumption of spices (and may be other spices) while conducting further surveys by the Government agencies like Indian Council of Medical Research (ICMR).

The traditional medicine system of India, Ayurveda, understands diet as ‘supreme medicine’.[30] As per its principles, a balance between different components of diet imparts ability to prevent and resist diseases. The spices mentioned in this study are included in many medicinal formulations in Ayurveda texts too. In the wake of COVID-19 pandemic, the Ministry of AYUSH that deals with traditional medicinal systems of India had published preventive remedies against this infection. Those guidelines had mentioned use of spices in form of turmeric milk and AYUSH kwatha consisting of ginger and black pepper.[31] Shankar Gautam et al have reviewed and justified role of each of these herbs in prevention of COVID-19.[32] Also, the source data does not take into account variations in spice usage according to season, change in diet during COVID-19 second wave, influence of AYUSH guidelines etc. Owing to these factors, it is possible that the consumption patterns of spices might have changed during subsequent second and third waves of COVID-19. It would be interesting if efforts are made to collect such data during next NSSO or other surveys being carried out for monitoring of COVID-19 situation and after too.

## Conclusion

The findings from this study certainly provide a basic framework and understanding. Further studies can be undertaken to know if spices specifically help in managing the course of disease. Their probable roles as adjuvant to treatment can also be explored. Should similar scenario occur in future, these findings can provide some vital base to act upon in adjuvant management. As the unspecified and under-explored ‘Other spices’ category showed promising correlation, more attention needs to be given to them too, along with mostly studied spices like ginger and turmeric.

## Data Availability

All data produced are available online at https://catalog.ihsn.org/catalog/3281/study-description and https://www.mohfw.gov.in/

## Author Contributions

The concept of the manuscript and search strategy was planned by SB. Literature searches and analysis was done by VB with key inputs from SB. The initial drafting of manuscript was done by VB and it was refined and finalized by SB.

## Ethics declarations

Not Applicable

## Acknowledgements

The authors are grateful to Dr DBA Narayana, Eminent Pharma Consultant, Bangalore for his guidance throughout drafting this manuscript. The authors also extend thanks to Dr Vinay Pawar, Mumbai for helping in statistical analysis. We also thank Dr. Ketki Wagh for her timely intellectual inputs.

## References

[1] Sarkar A, Chakrabarti AK, Dutta S. Covid-19 Infection in India: A Comparative Analysis of the Second Wave with the First Wave. Pathogens. 2021; 10(9):1222. https://doi.org/10.3390/pathogens10091222

[2] WHO Coronavirus Disease (COVID-19) Dashboard, Situation by Country, Territory & Area, https://covid19.who.int/table, x(Accessed September 3,2020)

[3] WHO-Coronavirus Dashboard-https://covid19.who.int/, (Accessed October 10,2020)

[4] COVID-19 State wise Status, https://www.mohfw.gov.in/,, (Accessed September 3,2020)

[5] Pere Domingo, Isabel Mura, Virginia Pomara, Hector Corominas, Jordi Casademont, Natividad de Benito, The four horsemen of a viral Apocalypse: The pathogenesis of SARS-CoV-2 infection (COVID-19), E-BioMedicine 58 (2020) 102887, https://doi.org/10.1016/j.ebiom.2020.102887

[6] Christianne de Faria Coelho-Ravagnani, Flavia Campos Corgosinho, Fabiane La Flor Ziegler Sanches, Carla Marques Maia Prado, Alessandro Laviano, João Felipe Mota, Dietary recommendations during the COVID-19 pandemic, Nutrition Reviews, uaa067, https://doi.org/10.1093/nutrit/nuaa067

[7] Acharya Charaka, Charaka Samhita, Description of HaritaVarga & Aharyogi Varga, sutrasthana chapter 27, verses 166-177 & 286-308, Chaukhamba orientalia, Varanasi, 2001

[8] Rajagopal K. Byran G, Jupudi S, Vadivelan R. Activity of phytochemical constituents of black pepper, ginger, and garlic against coronavirus (COVID □19): An in-silico approach. Int J Health Allied Sci 2020;9: S43–50. DOI: 10.4103/ijhas.IJHAS_55_20

[9] Vivek Kumar Soni, Arundhati Mehta, Yashwant Kumar Ratre, Atul Kumar Tiwari, Ajay Amit, Rajat Pratap Singh et al, Curcumin, a traditional spice component, can hold the promise against COVID-19?, European Journal of Pharmacology 886 (2020) 173551, https://doi.org/10.1016/j.ejphar.2020.173551

[10] Umesh, Debanjan Kundu, Chandrabose Selvaraj, Sanjeev Kumar Singh & Vikash Kumar Dubey, Identification of new anti-nCoV drug chemical compounds from Indian spices exploiting SARS-CoV-2 main protease as target, Journal of Biomolecular Structure and Dynamics, 2020, DOI: 10.1080/07391102.2020.1763202

[11] SpiceRx: an integrated resource for the health impacts of culinary spices and herbs, Rakhi Nk, Rudraksh Tuwani, Neelansh Garg, Jagriti Mukherjee, Ganesh Bagler bioRxiv 273599; DOI: https://doi.org/10.1101/273599

[12] Yehya Elsayed and Naveed Ahmed Khan, ACS Chemical Neuroscience 2020 11 (12), 1696–1698 DOI: 10.1021/acschemneuro.0c00239

[13] Nations within a nation: variations in epidemiological transition across the states of India, 1990–2016 in the Global Burden of Disease Study, L Dandona, R Dandona, GA Kumar, DK Shukla, VK Paul, K Balakrishnan, The Lancet 390 (10111), 2437–2460

[14] National Sample Survey Organization (NSSO). Household Consumption of Various Goods and Services in India 2011-12. New Delhi, India: Ministry of Statistics and Programme Implementation, Government of India; 2014. https://catalog.ihsn.org/catalog/3281/study-description

[15] Census data of states and UT-Population,2011-https://www.censusindia.gov.in/2011census/PCA/PCA_Highlights/pca_highlights_file/India/Chapter-1.pdf, (Accessed October 10,2020)

[16] Tay M.Z., Poh C.M., Rénia L., McAry P.A., Ng L.F.P. The trinity of COVID-19: immunity, inflammation and intervention. Nat Rev Immunol, 2020, 20, 363–374. https://doi.org/10.1038/s41577-020-0311-8

[17] Nogueira de Melo, G.A., Grespan, R., Fonseca, J.P. et al. Inhibitory effects of ginger (Zingiber officinale Roscoe) essential oil on leukocyte migration in vivo and in vitro. J Nat Med, 2011, 65, 241– 246. https://doi.org/10.1007/s11418-010-0479-5

[18] Manel Amri, Chafia Touil-Boukoffa, in vitro anti-hydatic and immunomodulatory effects of ginger and [6]-gingerol, Asian Pacific Journal of Tropical Medicine 2016; 9(8): 749–756, http://dx.doi.org/10.1016/j.apjtm.2016.06.013

[19] Rodrigo Arreola, Saray Quintero-Fabián, Rocío Ivette López-Roa, Enrique Octavio Flores-Gutiérrez, Juan Pablo Reyes-Grajeda, Lucrecia Carrera-Quintanar. Immunomodulation and Anti-Inflammatory Effects of Garlic Compounds, Immunomodulation and anti-inflammatory effects of garlic compounds. Journal of immunology research, 2015, 401630. https://doi.org/10.1155/2015/401630

[20] Krishnapura Srinivasan, Cumin (Cuminum cyminum) and black cumin (Nigella sativa) seeds: traditional uses, chemical constituents, and nutraceutical effects, Food Quality and Safety, 2018, 2(1),1–16, https://doi.org/10.1093/fqsafe/fyx031

[21] B. Laribi, K. Kouki, M. M’Hamdi, T. Bettaieb, Coriander (Coriandrum sativum L.) and its bioactive constituents, Fitoterapia,2015, 103, pp. 9–26, doi-10.1016/j.fitote.2015.03.012

[22] Boroumand N, Samarghandian S, Hashemy SI. Immunomodulatory, anti-inflammatory, and antioxidant effects of curcumin. J Herbmed Pharmacol. 2018;7(4):211–219. DOI: 10.15171/jhp.2018.33.

[23] Kottarapat Jeena, Vijayasteltar B. Liju, N.P. Umadevi &Ramadasan Kuttan, Antioxidant, Anti-inflammatory and Antinociceptive Properties of Black Pepper Essential Oil (Piper nigrum Linn), Journal of Essential Oil-Bearing Plants, 17:1, 1–12, DOI: 10.1080/0972060X.2013.831562

[24] Amin F. Majdalawieh and Ronald I. Carr. In Vitro Investigation of the Potential Immunomodulatory and Anti-Cancer Activities of Black Pepper (Piper nigrum) and Cardamom (Elettaria cardamomum), Journal of Medicinal Food. 2010.371–381.http://doi.org/10.1089/jmf.2009.1131

[25] Pundir, *Reena, Rani, R., Tyagi, S., & Pundir, P. Advance Review on Nutritional Phytochemical, Pharmacological and Antimicrobial Properties Of Chili, International Journal of Ayurveda and Pharma Research, 2016; 4(4). Retrieved from http://ijaprs.com/index.php/ijapr/article/view/332

[26] Paula FS, Kabeya LM, Kanashiro A, de Figueiredo AS, Azzolini AE, Uyemura SA, Lucisano-Valim YM. Modulation of human neutrophil oxidative metabolism and degranulation by extract of Tamarindus indica L. fruit pulp. Food Chem Toxicol. 2009 Jan;47(1):163–70. DOI: 10.1016/j.fct.2008.10.023. Epub 2008 Nov 3. PMID: 19022329.

[27] Spices under the purview of the Spices Board, https://www.indianspices.com/sites/default/files/spices%20list%20under%20purview%20of%20spices%20board.pdf,, (Accessed September 3,2020)

[28] Lakshadweep Cuisine: The Sea on a Plate, https://indianculture.gov.in/food-and-culture/south/lakshadweep-cuisine-sea-plate (Accessed May 4, 2022)

[29] Consumer spend survey won’t be released: Ministry-The Economic Times, https://economictimes.indiatimes.com/news/economy/indicators/consumer-expenditure-survey-not-to-be-released-due-to-data-quality-issues-government/articleshow/72074837.cms,, (Accessed October 12,2020)

[30] Acharya Kashyapa. Kashyapasamhita. Edited by P.V. Tewari.1st Edition. Varanasi: Chaukambha Vishwabharati;1996. Khilasthana, 4^th^ chapter, verses 4-6, page-468

[31] Ministry of AYUSH Ayurveda’s immunity boosting measures for self-care during COVID 19 crisis, https://www.ayush.gov.in/docs/123.pdf,(Accessed October 3,2020)

[32] Shankar Gautam, Arun Gautam, Sahanshila Chhetri, Urza Bhattarai, Immunity against COVID-19: Potential role of Ayush Kwath, J Ayurveda Integr Med, https://doi.org/10.1016/j.jaim.2020.08.003 [Ahead of Print]

